# Examining the relationship between plasma pTau181 and cognitive decline, structural brain integrity, and biological ageing in midlife

**DOI:** 10.1101/2025.04.09.25325556

**Authors:** Ashleigh Barrett-Young, Erin E. Cawston, Brigid Ryan, Wickliffe C. Abraham, Antony Ambler, Tim Anderson, Kirsten Cheyne, Elizabeth Goodin, Sean Hogan, Renate M. Houts, David Ireland, Annchen R. Knodt, Jesse Kokaua, Tracy R. Melzer, Sandhya Ramrakha, Karen Sugden, Benjamin Williams, Phillipa Wilson, Avshalom Caspi, Ahmad R. Hariri, Terrie E. Moffitt, Richie Poulton, Reremoana Theodore

**Affiliations:** Dunedin Multidisciplinary Health & Development Research Unit, Department of Psychology, University of Otago, Dunedin, New Zealand; Centre for Brain Research, University of Auckland, Auckland, New Zealand; Department of Pharmacology and Clinical Pharmacology, University of Auckland, Auckland, New Zealand; Brain Health Research Centre, University of Otago, Dunedin, New Zealand; Department of Medicine, University of Otago, Christchurch, New Zealand; New Zealand Brain Research Institute, Christchurch, New Zealand; Department of Psychology and Neuroscience, Duke University, North Carolina, USA; Va’a o Tautai Centre for Pacific Health, University of Otago, Dunedin, New Zealand; Te Kura Mahi ā-Hirikapo | School of Psychology, Speech and Hearing, University of Canterbury, Christchurch, New Zealand; Pacific Radiology Canterbury, Christchurch, New Zealand; Social, Genetic and Developmental Psychiatry Centre, Institute of Psychiatry, Psychology and Neuroscience, King’s College London, London, UK

**Keywords:** Alzheimer disease, dementia, biomarkers, blood biomarkers, diagnosis, phosphorylated tau

## Abstract

**INTRODUCTION:** Although plasma pTau181 has been shown to accurately discriminate patients with Alzheimer’s disease from healthy older adults, its utility as a preclinical biomarker in middle-aged community-based cohorts is unclear.

**METHODS:** Participants were members of the Dunedin Multidisciplinary Health and Development Study, a longitudinal study of 1037 people born in New Zealand in 1972-1973. Plasma pTau181, MRI-based brain structure, and DunedinPACE (an epigenetic biomarker of biological ageing) were measured at age 45; cognition was measured in childhood and age 45.

**RESULTS:** We observed a wide range of pTau181 concentrations in our same-aged sample (n=856; M=13.6pg/mL, SD=9.1pg/mL). Males had significantly higher pTau181 concentrations than females. No statistically significant associations were observed with cognitive decline, lower structural brain integrity, or accelerated biological ageing.

**DISCUSSION:** In this midlife cohort, wide variation in pTau181 concentrations was present by age 45, but was not associated with patterns of AD-risk in cognition, brain structure, or biological ageing.

**Research in context:** *Systematic review:* Authors reviewed the literature using PubMed and Web of Science databases. While research on plasma biomarkers of AD has largely focused on older people with mild cognitive impairment or AD, there are few studies of plasma biomarkers among general middle-aged populations. Given the potential utility of plasma biomarkers of AD such as pTau181 in early screening for disease risk, examining the concentrations of pTau181 among a younger cohort free of dementia is important for possible future clinical implementation.

*Interpretation:* Plasma pTau181 concentrations varied widely among our same-aged sample, yet higher pTau181 was not associated with cognitive decline, lower MRI-estimated structural brain integrity, or accelerated biological ageing. These findings indicate that variability in pTau181 exists in middle-age, but independently of other AD risk factors.

*Future directions:* Understanding how variability in pTau181 concentrations in midlife may predict later AD and to what extent this is distinct from other risk factors is important for shaping the translational pathway from lab to clinic.

## 1. Background

Blood-based biomarkers of Alzheimer’s disease (AD) are increasingly appealing as a low-cost, minimally-invasive part of the AD diagnostic pathway. However, these biomarkers are under-characterised in the general population, particularly in middle-aged individuals without dementia.^1^ AD has a long preclinical phase, with pathology accumulating over 10-30 years prior to diagnosis.^2^ Increasingly, the focus of AD treatments is on disrupting AD pathology progression in the preclinical stage, as pharmaceutical treatments have limited efficacy in preserving or restoring cognitive function in more advanced stages of AD.^3^ Lifestyle interventions are also likely to be more effective at mitigating or delaying AD when implemented earlier in the lifecourse, taking a more preventive approach.^4^ Thus, biomarkers that can reliably detect the earliest indications of AD pathology are critically important to the future implementation of AD treatments.

Blood-based AD biomarkers reflect the pathological hallmarks of AD, including hyperphosphorylated tau that in the brain forms neurofibrillary tangles and is highly correlated with amyloid-β (Aβ) pathology.^5^ Elevated plasma concentrations of tau phosphorylated at threonine-181 (pTau181) are specific to AD and discriminate from other tauopathies.^6–9^ pTau181 concentrations increase with worsening AD symptoms, indicating that changes in plasma concentrations may be good for monitoring disease progression.^10,11^ Higher plasma pTau181 concentrations may predict progression to AD in cognitively unimpaired individuals^12^ and may be increased in Aβ-positive compared to Aβ-negative young adults,^13^ suggesting it may be a potential biomarker for the earliest stages of AD. However, while pTau181 is highly correlated with Aβ burden, the presence or extent of Aβ is not necessarily deterministic for AD.^14^

It is not currently known how long before clinical presentation of AD differences in pTau181 concentrations can be measured.^15^ Most studies of plasma biomarkers of AD have been conducted in older-aged samples; few have characterised the range of plasma pTau181 in middle-aged population cohorts. In the ARIC study, plasma pTau181 in midlife (mean age 58 years) was associated with incident all-cause dementia later in life (mean follow up 7.4 years), as were increases in plasma pTau181 from midlife to later life.^16^ Some previous findings indicate that pTau181 may only begin increasing later in life. Cooper et al. (2023) found that pTau181 concentrations were lower in people aged under 60 than those 60 or older, although pTau181 did appear to begin increasing in concentration before age 60.^17^ Similarly, when combining multiple indicators of AD risk, Gebre et al. (2024) found that pTau181 was an important predictor for older individuals but less so for younger (<65 years) individuals.^18^

Assessing population-representative samples of middle-aged individuals free from dementia is critical to understanding the temporal nature of plasma pTau181. The prognostic value of pTau181, like most putative AD-risk biomarkers, in real-world populations rather than highly selective research samples is an important step towards the translation of biomarkers to clinical care.^19^ In the Dunedin Study, we have previously shown that MRI-based measures of worse brain structural integrity, accelerated biological ageing, and greater cognitive decline are interrelated by age 45 and predictive of risk for AD and related dementias.^20^ These measures have also been shown to predict dementia in older cohorts.^21^ Should pTau181 be associated with other indicators of AD risk, it would suggest that plasma pTau181 may reflect similar pathological processes to those associated with risk as measured by MRI, accelerated biological ageing, or sensitive cognitive tests, and consequently could be a good biomarker for preclinical AD. Conversely, if pTau181 shows no association with such measures, it would suggest that pTau181 independently shapes AD risk in midlife.

The objectives of this study were to first quantify plasma pTau181 in a middle-aged, population-based cohort without dementia. Secondly, we aimed to examine whether plasma pTau181 was associated with AD risk features of cognition, structural brain integrity, and biological ageing. We hypothesized that higher pTau181 at age 45 would be associated with greater cognitive impairment and decline, worse structural brain integrity, and accelerated biological ageing.^21^

## 2. Method

### 2.1 Participants

Participants are members of the Dunedin Multidisciplinary Health and Development Study (‘Dunedin Study’), a longitudinal investigation of health and behaviour in a population-representative birth cohort of 1,037 individuals (91% of eligible births; 52% male) born between 1 April 1972 and 31 March 1973 in Dunedin, New Zealand (NZ). The longitudinal study was established at age 3 years based on residence in the province.^22,23^ Assessments were conducted at birth and at ages 3, 5, 7, 9, 11, 13, 15, 18, 21, 26, 32, 38, and most recently at age 45, when 94% of the 997 participants still alive took part. Each study member was brought to the research unit for a day of interviews and examinations. MRI scans were completed for 93% of eligible age-45 participants. The cohort represents the full range of socioeconomic status on NZ’s South Island, and as adults match the NZ National Health and Nutrition Survey on adult health indicators (e.g., body mass index (BMI), smoking, physical activity, general practitioner visits)^24^ and matches the NZ Census on educational attainment.^25^ None of the analytic sample reported having been diagnosed with AD or Down Syndrome. Study participants are primarily of New Zealand European ethnicity; 8.6% reported Māori ethnicity at age 45. Written informed consent was obtained from participants, and the study was approved by the New Zealand Health and Disability Ethics Committee (NZ-HDEC).

### 2.2 Plasma pTau181

Phlebotomy at age 45 was performed by Southern Community Laboratories in the late afternoon for all participants. Participants did not fast. Whole blood was collected in 10mL K_2_EDTA vacutainer tubes, centrifuged within 2 hours of collection (1300 x *g* for 10 minutes at room temperature), and plasma was aliquoted into polypropylene screw-top cryotubes that were immediately moved to ultra-low temperature (-80°C) freezers at the University of Otago. Plasma was collected in 2017-2019 and stored frozen until January 2023, when samples underwent single thawing for 2 minutes in a 37°C water bath and were pipetted up and down 3 times per sample, aliquoted, and immediately frozen prior to being shipped on dry ice to the University of Auckland for analysis. Anonymised aliquots of EDTA plasma were measured in duplicate using a Simoa SR-X platform at the University of Auckland. The immunoassaying kits used were pTau-181 Advantage V2.1 kits (QTX-104111; Quanterix), all from the same lot (503680). Lipemic samples were noted. Quality control (QC) materials supplied with the kits were analysed in duplicate at the start and end of each plate to assess precision. The mean concentration, repeatability and intermediate precision for the low and high QC were 37.8pg/mL, 8.4%, 14.2% and 568.5pg/mL, 8.8%, 8.8% respectively. Samples were diluted 1:4 per manufacturer’s recommendations. If a sample did not return a value in either duplicate, the assay was repeated. All samples that returned a value were retained; data from all repeated samples were averaged. Samples below the lower limit of detection were imputed at 0 (LLOD = 3.3pg/mL). The functional lower limit of quantification (LLOQ) was calculated as 12.44pg/mL; samples below the LLOQ were retained. N=867 Study members had samples available for assaying; of these, n=11 did not return any data from assaying. Study members who had chronic kidney disease (CKD; n=2) were included in the distributional analysis (Figure 1) but omitted from all other analyses. N=854 Study members were included in final analysis.

**Figure 1.**
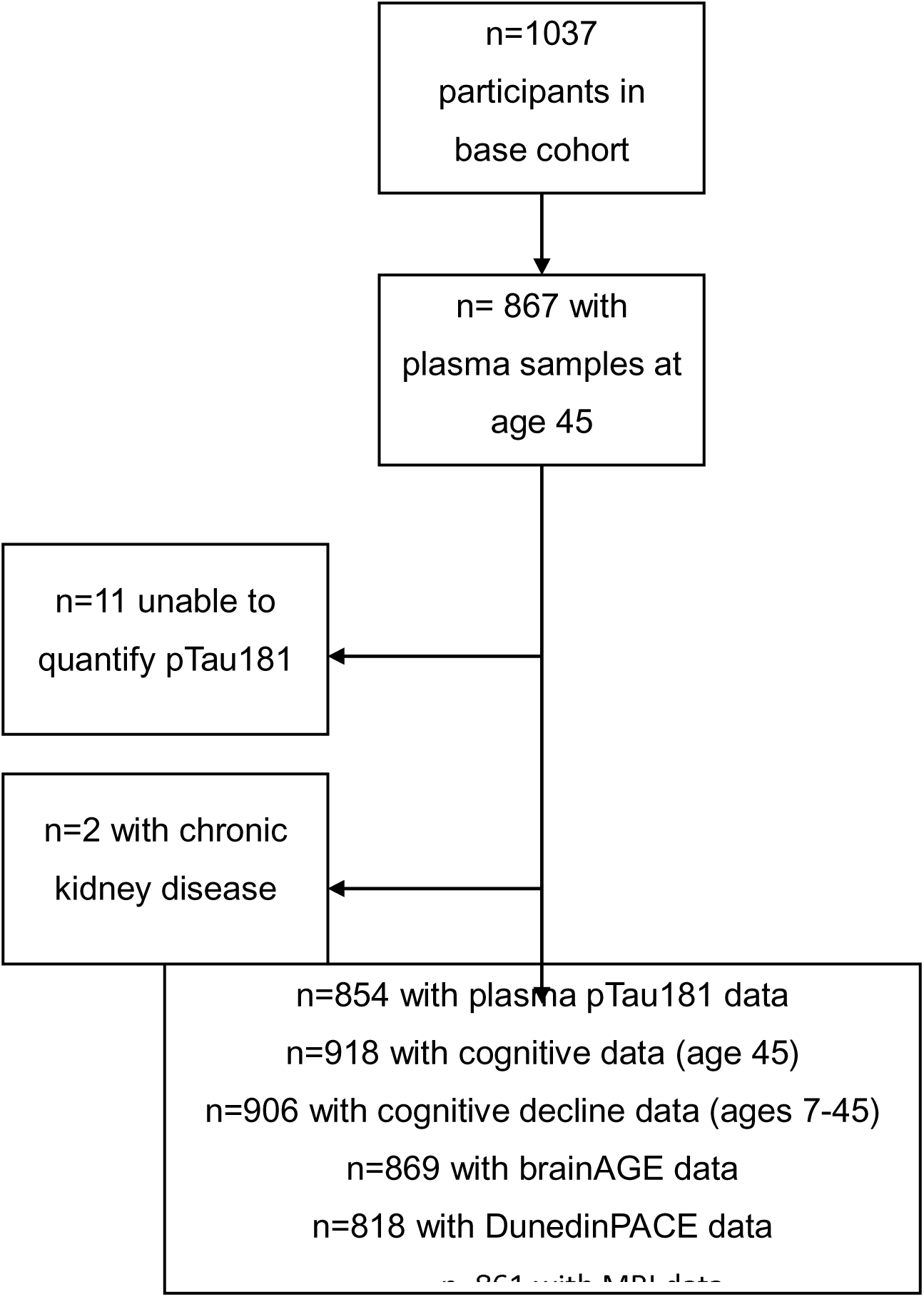
Participant flow chart.

### 2.7 Data analysis

Analyses were conducted in R and Stata between November 2023-December 2024. Descriptive statistics for the unadjusted pTau181 concentrations are presented. Non-parametric tests were used to compare groups (Wilcoxon rank sum test for biological sex and Kruskal-Wallis for number of APOEε4 alleles). For regression analyses, pTau181 concentration was log transformed to meet assumptions of normality. For each analysis, pTau181 was used to predict the variable of interest in a sex-adjusted model, followed by model adjusted for sex and BMI, then a sex- and APOEε4-adjusted model. First, pTau181 concentration was used to predict global cognitive decline on a continuous scale. For post-hoc analyses, we categorised participants according to cognitive decline since childhood; first, those with greater than 1 standard deviation (>15 IQ points) decline; and then those with greater than 2 standard deviations (>30 IQ points) decline. Logistic regression models were constructed to predict whether pTau181 concentration, adjusted for sex, predicted each category of cognitive decline.

To test for associations between pTau181 and brain structural integrity, linear regression models were constructed where pTau181 was entered as a predictor of 1) brainAGE; 2) WMH volume, which was log transformed as the data were non-normal; 3) mean cortical thickness and average surface area; 4) mean fractional anisotropy; and 5) subcortical grey matter volumes. Finally, to explore the distribution patterns of associations, parcel-wise analyses of cortical thickness and surface area were conducted. In these post-hoc analyses, we ran linear regressions using pTau181 to predict the surface area and thickness of 360 cortical parcels.^34^ For further post-hoc analysis, we categorised participants with the top 5% of brainAGE scores (indicating the oldest brains), and used logistic regression, adjusted for sex, to determine whether pTau181 predicted having the top 5% brainAGE score. Finally, we tested for an association between pTau181 concentration and DunedinPACE using linear regression.

We corrected for multiple comparisons across the subcortical and cortical parcel-wise analyses by using a false discovery rate (FDR) procedure;^35^ for analyses of all other variables we used an alpha level of .05. All tests were one-sided. Sex was included as a covariate in all regression analyses. Analyses with subcortical grey matter volumes were repeated controlling for total brain volume, which tests relative size of a region rather than absolute size.^36^ The premise and analysis plan for this study was preregistered in July 2023 (https://dunedinstudy.otago.ac.nz/files/1692931368_Barrett-Young_pTau181inDunedinStudy_CP_24-07-2023.pdf). Analyses were checked for reproducibility by a statistician (JK), who used the manuscript to verify the statistical code and applied it to a fresh copy of the dataset.

## 3. Results

Collection of plasma samples occurred at the Age 45 assessment (M=45.3 years, SD=0.55 years), between April 2017 and April 2019. The analytic dataset included Study members with plasma pTau181 data available (N=856; female 49.3%, male 50.7%; **Figure 1**). Participants with chronic kidney disease (CKD) had exceptionally high concentrations of pTau181 (>100pg/mL); these Study members (n=2) were included in the distribution of pTau181 (**Figure 2**) but removed from regression analyses, leaving n=854.

**Figure 2.**
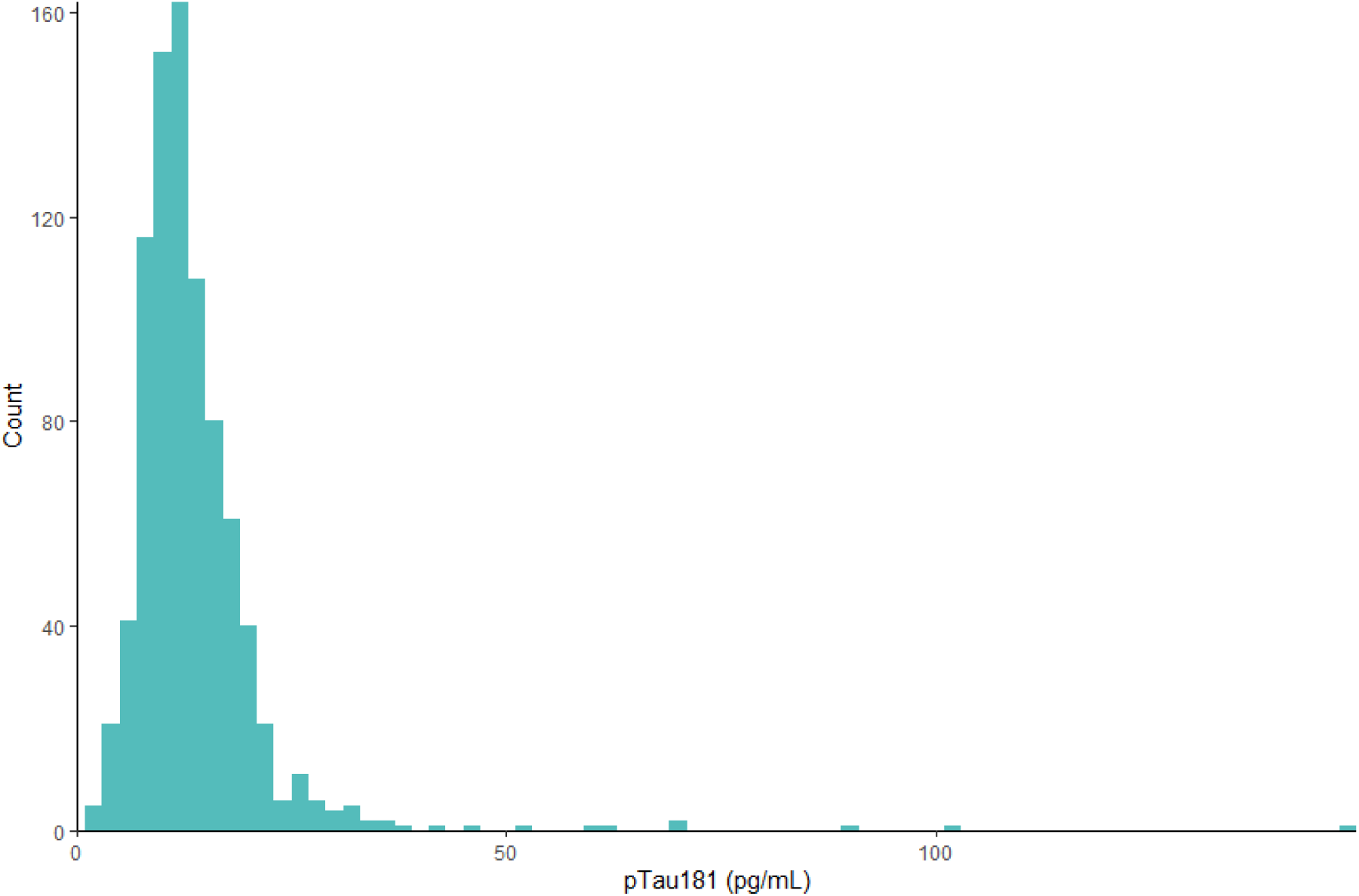
Distribution of pTau181 concentrations among 45-year-olds.

A wide distribution of pTau181 concentrations was observed, with n=2 under the lower limit of detection (M=13.6pg/mL, SD=9.1; **Figure 2).** Once participants with CKD were removed, the maximum value was 90.6pg/mL (M=13.4pg/mL, SD=7.2).

Males had higher pTau181 concentrations than females on average (M(SD)_male_=14.7(8.5); M(SD)_female_=12.0(5.4), *W*=71179, *p*<.001; **Figure 3**). A post-hoc two-way ANOVA was conducted to determine whether the sex difference was explained by a difference in BMI between the sexes; while the main effect of sex remained significant (*F*(1, 841)=19.05, *p*<.001), neither the main effect of BMI (*F*(5, 841)=0.90, *p*=.48) nor the interaction of sex and BMI (*F*(4, 841)=0.08, *p*=.80) were significant. Thus, the sex difference observed was independent of BMI categories.

**Figure 3.**
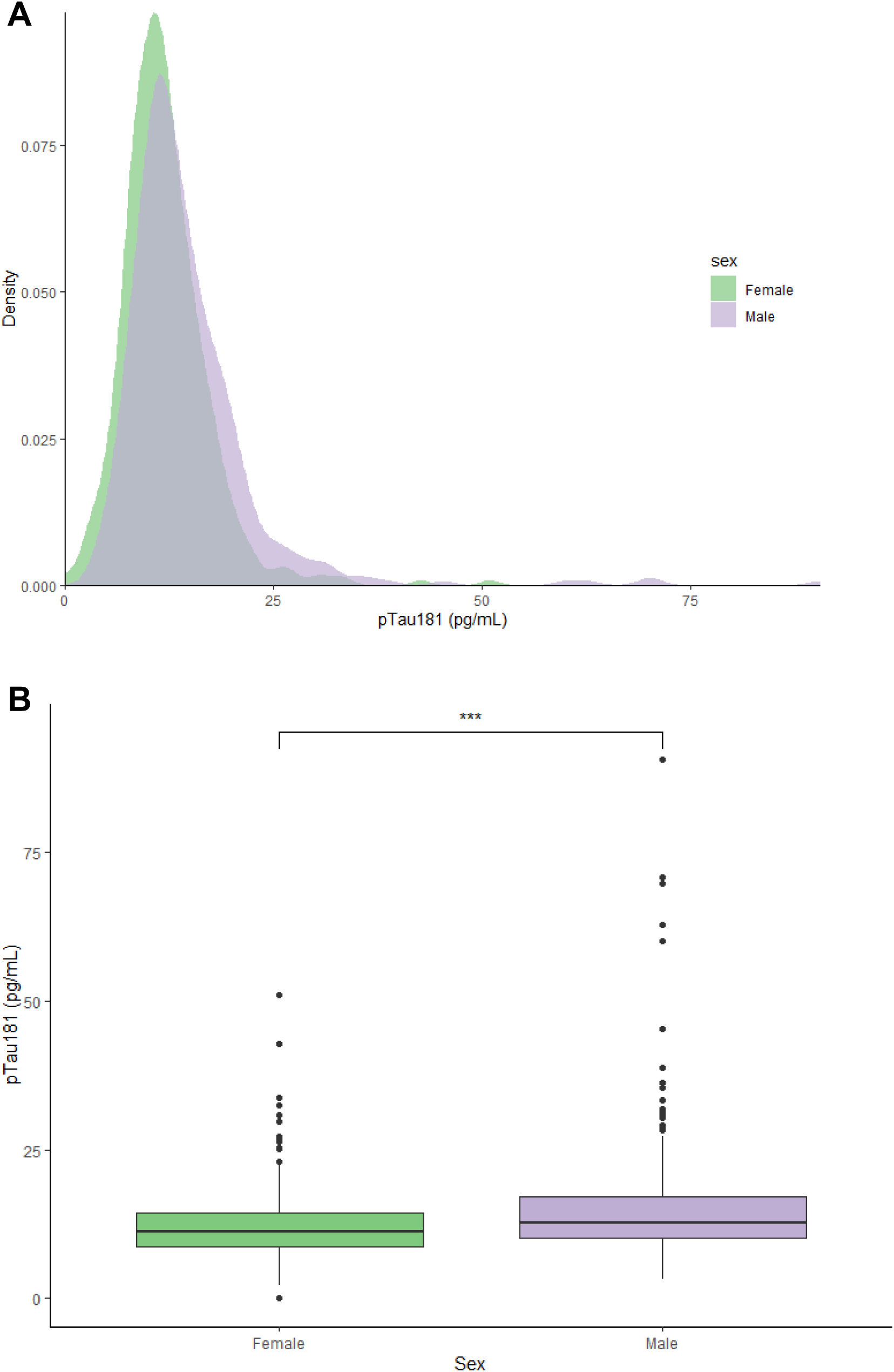
Sex differences in pTau181 concentrations

There were no statistically significant differences in pTau181 concentrations between participants with one or two copies of the APOEε4 allele or with those with none (*H*(2)=1.8, *p*=.40; **Figure 4**, **Table 1**).

**Figure 4.**
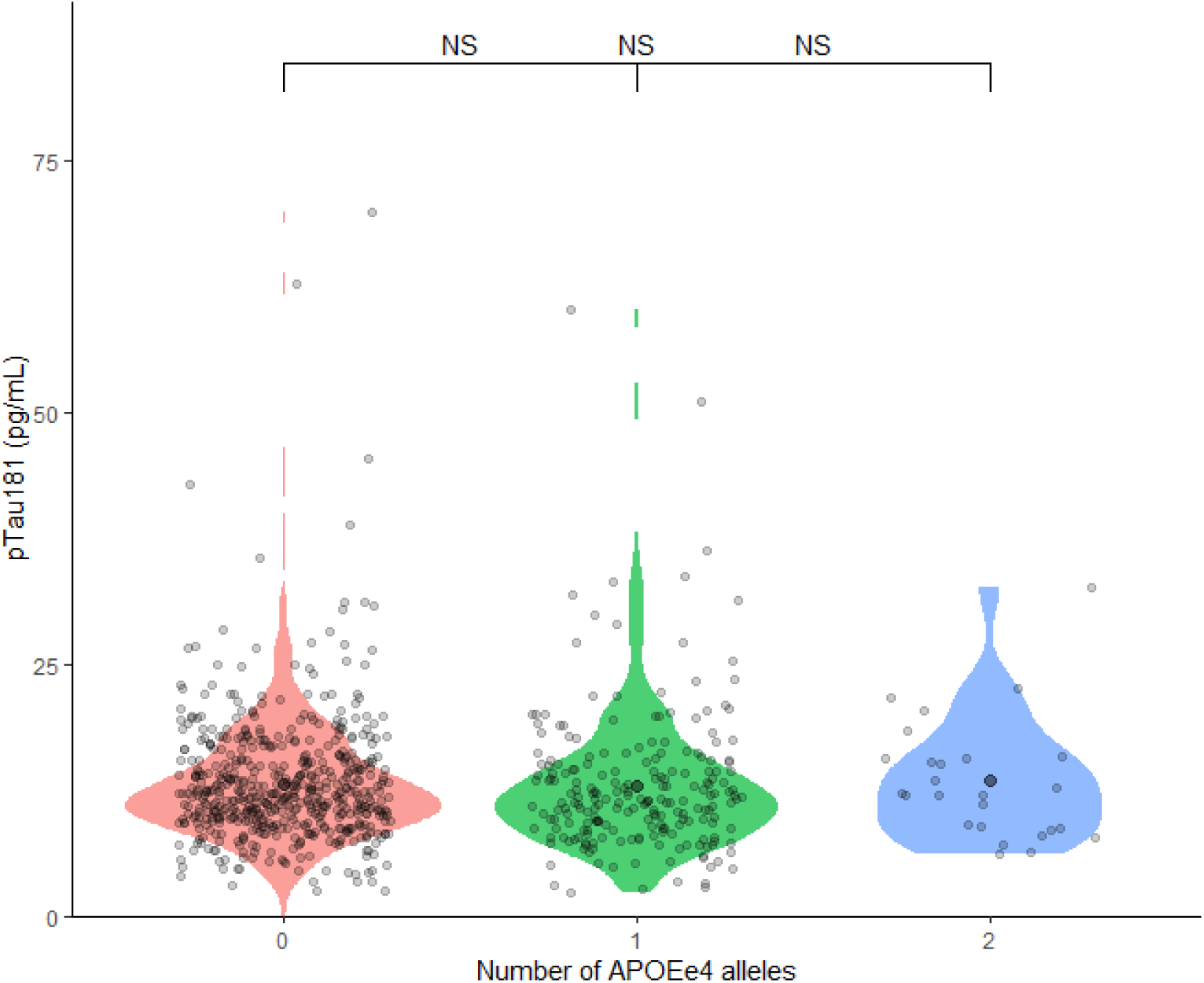
pTau181 concentrations by number of APOEε4 alleles

**Table 1.** Mean and standard deviation of plasma pTau181 concentrations (pg/mL) by number of APOEε4 alleles.

|  | N (%) | M | SD |
| --- | --- | --- | --- |
| None | 556 (69.5%) | 13.2 | 6.34 |
| One | 220 (27.4%) | 13.0 | 7.12 |
| Two | 26 (3.2%) | 13.5 | 6.05 |

We did not observe any associations between pTau181 and measures of cognitive impairment or decline; structural brain integrity including brainAGE, WMH volume, mean cortical thickness, average cortical surface area, mean fractional anisotropy, and subcortical grey matter volumes; or accelerated biological ageing (**Table 2**). Exploratory analyses with parcel-wise cortical thickness and surface area as well as tract-wise fractional anisotropy revealed no statistically significant associations after correction for multiple comparisons (**Supplementary materials**). In post-hoc analyses, we categorised participants according to cognitive decline (greater than 1 standard deviation decline in cognition since childhood [n=137; 16% of the analytic sample]; greater than 2 SD decline in cognition since childhood [n=15, 1.4% of the full cohort]). Logistic regression revealed that pTau181 did not predict having more than 1 SD decline in cognition (χ^2^(2)=3.6, *p*=.16; OR=0.77[95% CI: 0.51, 1.16], *p*=.21), nor did it predict greater than 2 SD decline in cognition (χ^2^(2)=2.5, *p*=.29; OR=0.46(95% CI: 0.17, 1.31), *p*=.15). In addition, we categorised participants with the oldest 5% of brainAGE scores; logistic regression showed that pTau181 did not predict being in the top 5% of oldest brainAGE scores (χ^2^(2)=3.3, *p*=.19; OR=0.82(95% CI: 0.41, 1.63), *p*=.57).

**Table 2.** Tests of associations between plasma pTau181 and measures of cognitive decline, structural brain integrity, and biological ageing.

|  | Model 1 (pTau181 + sex) |  |  |  | Model 2 (pTau181 + sex + BMI) |  |  |  | Model 3 (pTau181 + sex + APOEε4) |  |  |  |
| --- | --- | --- | --- | --- | --- | --- | --- | --- | --- | --- | --- | --- |
| | $\beta$ | Lower CI | Upper CI | $p$ | $\beta$ | Lower CI | Upper CI | $p$ | $\beta$ | Lower CI | Upper CI | $p$ |
| **Cognitive decline** | 0.04 | -0.03 | 0.10 | .32 | 0.03 | -0.03 | 0.10 | .33 | 0.03 | -0.04 | 0.10 | .41 |
| **brainAGE** | -0.04 | -0.11 | 0.03 | .25 | -0.04 | -0.11 | 0.03 | .25 | -0.07 | -0.14 | 0.00 | .06 |
| **Total surface area** | 0.01 | -0.05 | 0.07 | .72 | 0.01 | -0.05 | 0.07 | .72 | 0.03 | -0.03 | 0.09 | .27 |
| **Mean cortical thickness** | 0.00 | -0.07 | 0.07 | .89 | 0.00 | -0.07 | 0.07 | .92 | 0.04 | -0.03 | 0.11 | .30 |
| **Mean fractional anisotropy** | 0.00 | -0.07 | 0.07 | .91 | 0.00 | -0.06 | 0.07 | .89 | 0.05 | -0.03 | 0.12 | .21 |
| **White matter hyperintensity volume** | 0.01 | -0.06 | 0.08 | .87 | 0.01 | -0.06 | 0.08 | .86 | 0.01 | -0.06 | 0.09 | .75 |
| **DunedinPACE** | -0.04 | -0.11 | 0.03 | .30 | -0.05 | -0.11 | 0.02 | .14 | -0.04 | -0.11 | 0.03 | .26 |

*Note.* $\beta$ : standardised regression coefficient; CI: 95% confidence interval.

## 4. Discussion

We found a wide range of plasma pTau181 concentrations among our population-representative sample of 45-year-olds. Males had, on average, slightly higher pTau181 concentrations than females. Those with two APOEε4 alleles had a slightly higher mean pTau181 compared to those with one or no APOEε4 alleles; however, this difference was not statistically significant. There were no statistically significant associations with MRI-based measures of structural brain integrity, nor were there between pTau181 and either cognition or cognitive decline. Moreover, pTau181 concentrations were not significantly associated with biological ageing, measured by DunedinPACE. Overall, these findings suggest, while the range of plasma pTau181 concentrations among middle-aged adults is wide, this variability is not associated with AD risk-related differences in cognition, structural brain integrity, or biological ageing at this age. Although AD is more prevalent in older people,^37^ evidence suggests AD pathology begins to accumulate decades prior.^38,39^ Previous research in clinical populations has shown that plasma pTau181 is associated with Aβ burden, tauopathy consistent with AD, and cognitive impairment.^9,40,41^ Plasma pTau181 predicted future AD diagnosis and greater hippocampal atrophy among the Alzheimer’s Disease Neuroimaging Initiative cohort.^43^ Our findings may suggest that variability in pTau181 reflects normal variation in the middle-aged population and may not reflect AD risk in midlife. Alternatively, given findings from previous studies, another interpretation of our findings is that plasma pTau181 is already present in high concentrations in midlife and may be an early indicator of AD, but is not related to other established AD risk factors. Further research in other middle-aged population cohorts, combined with longitudinal follow up of these cohorts at later ages, is required to better understand the potential clinical utility of plasma pTau181 in midlife. Plasma pTau181 may be particularly sensitive to intra/inter-assay differences and variation depending on pre-analytic and analytic methods. Cooper at al. (2023) found that pTau181 was subject to cross-lot bias;^17^ we minimised intra-assay differences by using kits from the same lot, running all analyses on the same machine, and running samples in duplicate. Plasma pTau181 has been shown to be stable across up to three freeze-thaw cycles.^44^ The majority of the samples in this study underwent two freeze-thaw cycles (n=749); a minority (n=29) were freeze-thawed 4-5 times. Furthermore, there may be differences in the raw concentrations of plasma pTau181 reported by various studies due to methodological differences, including different assaying technologies used. In a study using the same device (Simoa SR-X) and immunoassaying kits (Quanterix pTau-181 Advantage V2.1) as the present study, Mohaupt et al. (2024) found plasma pTau181 concentrations among n=20 cognitively unimpaired individuals ranged from 5.33pg/mL to 78.2pg/mL (M=18.1pg/mL),;^45^ a similar range to that found in our study. In one study of n=16 cataract patients with normal cognition, plasma pTau181 range was 0.66-17.7pg/mL (M=3.2pg/mL);^46^ another study of n=65 cognitively normal older adults found a mean of 23.5pg/mL (SD=9.6pg/mL), though no range was reported.^47^ The wide ranges and variation in mean concentrations observed in these studies, in which the same equipment was used, could be due to low sample sizes that do not represent the full range of plasma pTau181 in the population. Thus, our study of n=854 individuals, none of whom have been diagnosed with AD, suggests the range of plasma pTau181 concentrations in the cognitively unimpaired midlife population is wide.

Females are diagnosed with AD at a higher rate than males, although it is unclear to what extent biological sex or gender differences contribute to this difference.^48^ Some studies have found females tend to have higher levels of CSF tau and tau-PET^49,50^ and greater tau tangle density at autopsy,^51^ although findings on plasma pTau181 are more mixed.^6,17,52^ One study found that even when females and males had similar levels of plasma pTau181, females tended to have worse phenotypic outcomes.^53^ A recent meta-analysis found that females accumulated tau faster than men, suggesting that, in those with high Aβ burden, tauopathy may be more aggressive and accumulate faster.^54^ Thus, while in our cohort males had higher pTau181 than females, the rate of accumulation and resulting phenotypic alterations may differ between the sexes later in life. It is possible that females have lower baseline tau but accumulate it faster; males may have higher baseline tau but accumulate tau more slowly. Interestingly, one of the few studies of plasma pTau181 in midlife also found that males had higher pTau181 concentrations than females.^55^ We intend to repeat plasma pTau181 measures in later assessments of the Dunedin Study to investigate this question further. In the revised criteria for AD diagnosis from the National Institutes on Aging and the Alzheimer’s Association (NIA-AA), CSF pTau181 is listed as a Core 1 biomarker, i.e., an early-changing biomarker of preclinical AD, though it is as yet unclear whether plasma pTau181 is sufficiently accurate to be included.^56^ While there is debate on the role of biomarkers in establishing a diagnosis of AD, particularly in the absence of cognitive symptoms,^57^ it is important that the variation of pTau181 in the population is understood. The data from our middle-aged cohort, none of whom had diagnosed AD, shows that the range of pTau181 in this age group is very wide and not yet associated with AD-risk-related variability in cognition, structural brain integrity, or biological ageing. Future assessments of this cohort will allow us to examine the progression of pTau181 concentrations across midlife and into later life, enabling us to examine if or when this biomarker begins to index preclinical AD. Blood biomarkers are ideal for initial screening in primary care as they are minimally invasive, low cost, and accessible in rural and underfunded areas.^58^ Therefore, it is imperative that plasma pTau181 is studied in population-based, unselected cohorts in natural settings. This study was conducted in a birth cohort that is representative of the general population of Aotearoa New Zealand on socioeconomic status and physical health. It has achieved exceptionally high retention rates (>90% at every adult assessment), so it is less subject to attrition or selection biases than many other studies. However, this also means that this sample is likely to include individuals who are experiencing cognitive decline due to other, non-AD-related reasons, including acquired brain injuries, pharmaceutical effects, substance and/or alcohol use, or other factors that may affect cognition. We were unable to account for every potential confounding factor in all analyses. Of potential comorbidities that may affect plasma pTau181, CKD has been clearly established as influencing pTau181 concentrations,^59–61^ although the direction of causality is not entirely clear.^62^ Notably, participants with clinician- and biomarker-confirmed CKD diagnoses recorded the highest pTau181 concentrations in this sample (>100pg/mL). BMI is negatively associated with pTau181 concentrations;^63^ however, adding BMI to analysis did not change our results.

Importantly, more research in population-based cohorts is needed to understand which factors affect pTau181 and should therefore be considered when interpreting biomarker results. The main limitation of this study is that our data are largely cross-sectional and only available at age 45, so we currently lack data at older ages that would allow us to test if plasma pTau181 at age 45 is an independent predictor of dementia at later ages. We found that plasma pTau181 can be measured at age 45 but does not relate to other AD risk factors at this age. One potential implication is that plasma pTau181 should not be used for early detection of dementia in young-middle-aged people; another implication is that, because pTau181 is measurable at age 45 but not related to other risk factors, it may be an independent predictor of dementia that could improve early identification. Without long-term follow-up into old age, we are currently unable to adjudicate between these two implications. Future assessment phases of this cohort will provide further insights.

According to the ATN model, biomarkers of Aβ, tauopathy, and neurodegeneration are used to classify AD stages.^56,64^ In the absence of markers of Aβ or neurodegeneration, we are currently unable to distinguish individuals in the Dunedin Study cohort who are Aβ-positive from those who are Aβ-negative and are therefore unable to conclusively determine whether any of our participants have preclinical AD. We plan to expand the panel of plasma biomarkers of AD at the next assessment (age 52). Thus, any cognitive decline and neurostructural abnormalities among the sample cannot be singularly attributed to preclinical AD and may be due to other neurocognitive disorders, such as dementia with Lewy bodies or frontotemporal dementia, long-term lead exposure,^65^ or some mental health disorders.^66^

## Conclusion

There is potential clinical utility of blood biomarkers of AD in the early part of the AD diagnostic pathway, and plasma pTau181 has emerged as one of the most promising candidates to identify AD pathology. In this middle-aged birth cohort without AD, we observed a wide range of pTau181 concentrations. In contrast with many studies of older participants, we found that males had higher concentrations of pTau181 than women. pTau181 concentrations were not associated with cognitive decline (measured from childhood to age 45), worse structural brain integrity, or biological ageing. Thus, AD risk associated with midlife variability in pTau181 may be independent of other known risk factors.

## Data Availability

All data produced in the present study are available upon reasonable request to the authors.

## Acknowledgements

We thank the Dunedin Study members, their families, and friends for their long-term involvement. The Dunedin Multidisciplinary Health and Development Research Unit is based at University of Otago within the Ngāi Tahu tribal area, whom we acknowledge as first peoples, tangata whenua (transl. people of this land). We thank Unit research staff and Study founder, Dr. Phil A. Silva. We would like to acknowledge the assistance of the Molecular Genomics Core at the Duke Molecular Physiology Institute, Duke University School of Medicine, for the generation of DNA data used in this project.

## Data availability statement

Proposed data-analysis projects from qualified scientists must have a concept paper describing the purpose of data access, IRB approval at the applicants’ university, and provision for secure data access. We offer secure access on the Otago and Duke campuses. These access requirements parallel those used by dbGap and the Health and Retirement Study.

## Abbreviations

Aβ: Amyloid beta
AD: Alzheimer’s disease
APOEε4: Apolipoprotein E isoform ε4
BMI: Body mass index
brainAGE: Brain age gap estimate
CKD: Chronic kidney disease
DunedinPACE: Dunedin Pace of Ageing Calculated from the Epigenome
EDTA: Ethylenediaminetetraacetic acid
FSIQ: Full scale IQ
GMV: Grey matter volumes
MRI: Magnetic resonance imaging
NFT: Neurofibrillary tangles
pTau181: phosphorylated tau at threonine 181
WMH: White matter hyperintensities

