## Supplementary materials for "Examining the relationship between plasma pTau181 and cognitive decline, structural brain integrity, and biological ageing in midlife"

in a middle-aged birth cohort

Ashleigh Barrett-Young, Erin E. Cawston, Brigid Ryan, Wickliffe C. Abraham, Antony Ambler, Tim Anderson, Kirsten Cheyne, Elizabeth Goodin, Sean Hogan, Renate M. Houts, David Ireland, Annchen R. Knodt, Jesse Kokaua, Tracy R. Melzer, Sandhya Ramrakha, Karen Sugden, Benjamin Williams, Phillipa Wilson, Avshalom Caspi, Ahmad R. Hariri, Terrie E. Moffitt, Richie Poulton, & Reremoana Theodore

Table of Contents

**Phase 45 Attrition Analysis**	**2**

**Magnetic Resonance Imaging**	**5**

Procedure	5

Image acquisition parameters	5

Image processing	5

Brain Age Gap Estimate	5

White matter hyperintensities	5

Subcortical grey matter volume	6

Parcel-wise cortical surface area and cortical thickness	6

Fractional anisotropy	6

References	6

**Supplementary Tables**	**8**

S1. Associations between pTau181 levels are age 45 and cognitive performance	8

S2. pTau181 and global MRI variables (relative values, controlled for total brain volume)	9

S3. pTau181 and grey matter volumes of subcortical structures (absolute values)	10

S4. pTau181 and grey matter volumes of subcortical structures (relative values, controlled for total brain volume)	11

### Phase 45 Attrition Analysis

We conducted an attrition analysis using childhood SES, childhood IQ, and history of psychopathology to determine whether participants in the Phase 45 data collection were representative of the original cohort. We report childhood SES and childhood IQ because they are known to be strong predictors of late-life health outcomes, as shown by many cohort studies from many nations. Childhood SES and childhood IQ separately predict health and social outcomes in adulthood, and these outcomes include physical functions, cognitive decline, mental health, inflammation, metabolic syndrome, disease incidence, dementia, mortality, and also neuroimaging-based, genomic, and epigenetic indicators of health.

Based on the literature, we report three groups: Study members who died before age 45 and thus could not have taken part in data collection, Study members who were alive and thus could take part, and Study members who actually did take part. We compared these three groups to the original birth cohort. The figures below show that the small group of study members who had died before age 45 had significantly lower mean childhood IQ on average as a group (*t* = 2.09, *p* = 0.04), and somewhat lower mean childhood SES, though these were not statistically significant. (Bars represent the range of values in the cohort.) Some of the early deaths were Dunedin Study members who had more disadvantages in their lives leading to poorer health and increased risk of early mortality. Study members who died of childhood diseases may have been already unwell at the time of IQ testing, which could have lowered their scores. However, cohort members who are still alive and cohort members who took part in data collection did not differ from the full original cohort on their mean childhood IQ and SES; they still represent population variation on these key health risk factors.

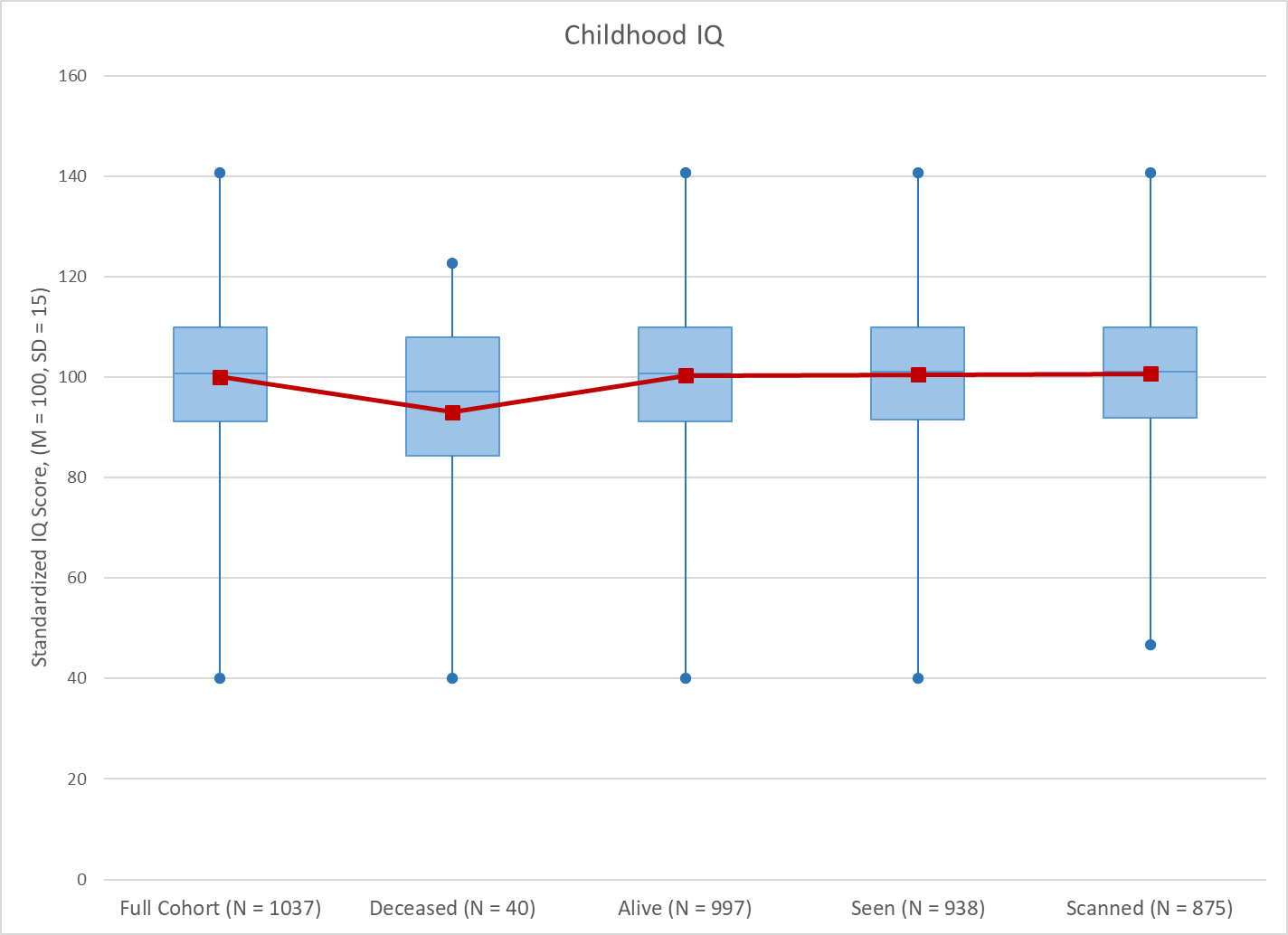

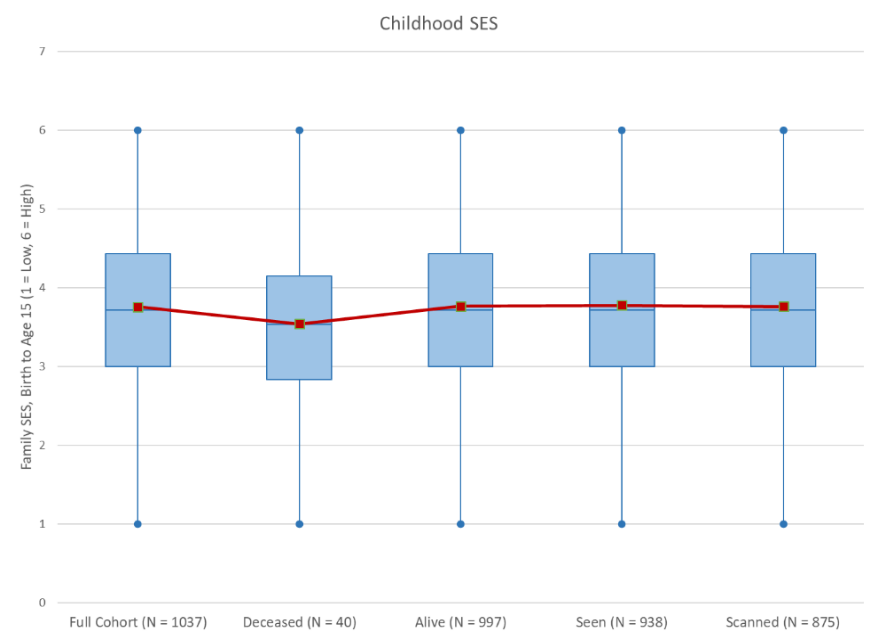

No significant differences in p-factor were found between the full cohort, those still alive, those seen at Phase 45 or those scanned at Phase 45. Those who were deceased by the Phase 45 data collection had significantly higher p-factor scores than those who were still alive (*t* = -2.86, *p* = 0.004).

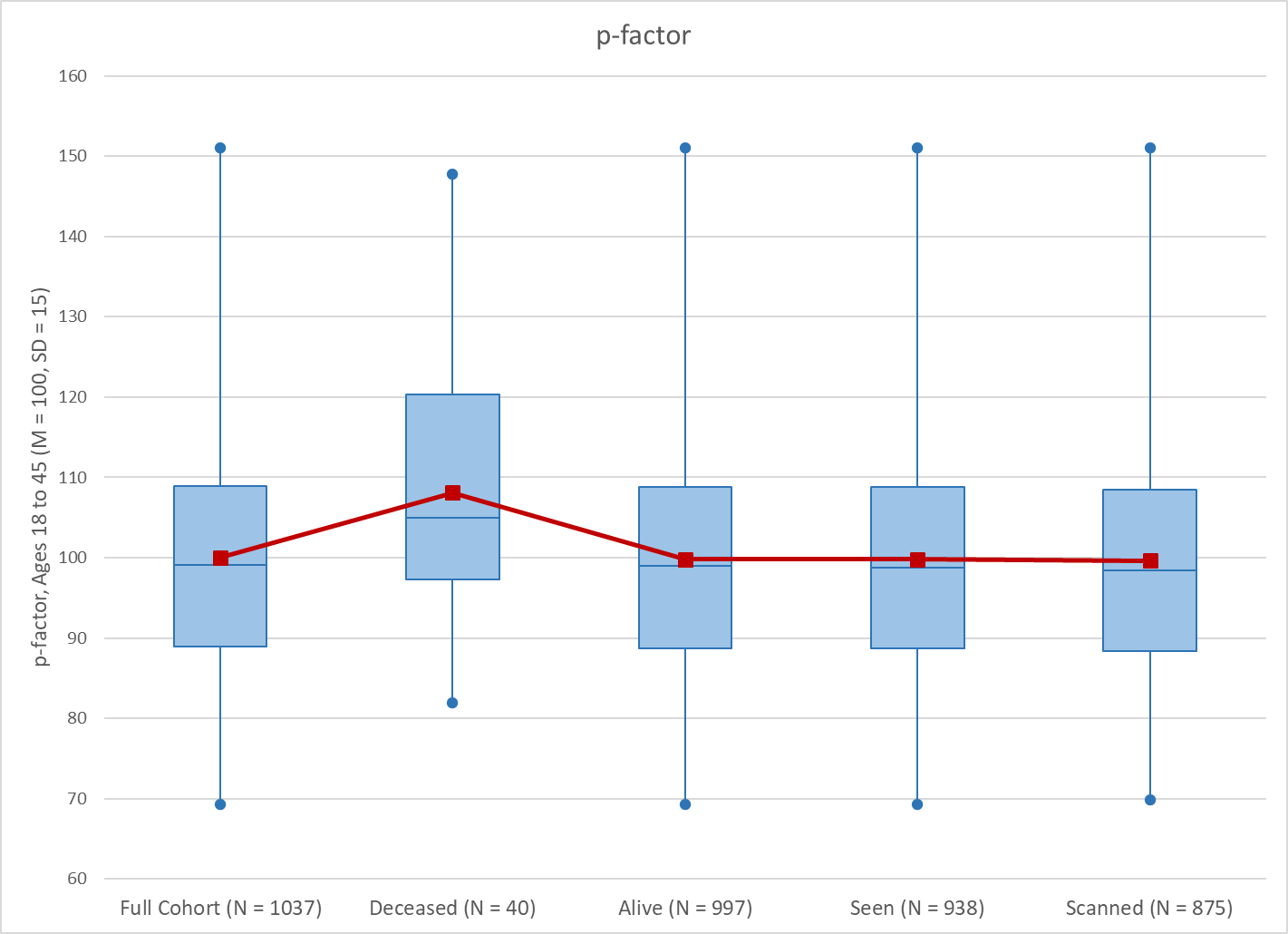

No significant differences were found between the full cohort, those deceased, those alive, those seen at Phase 45 or those scanned at Phase 45 on Adverse Childhood Events (ACEs).

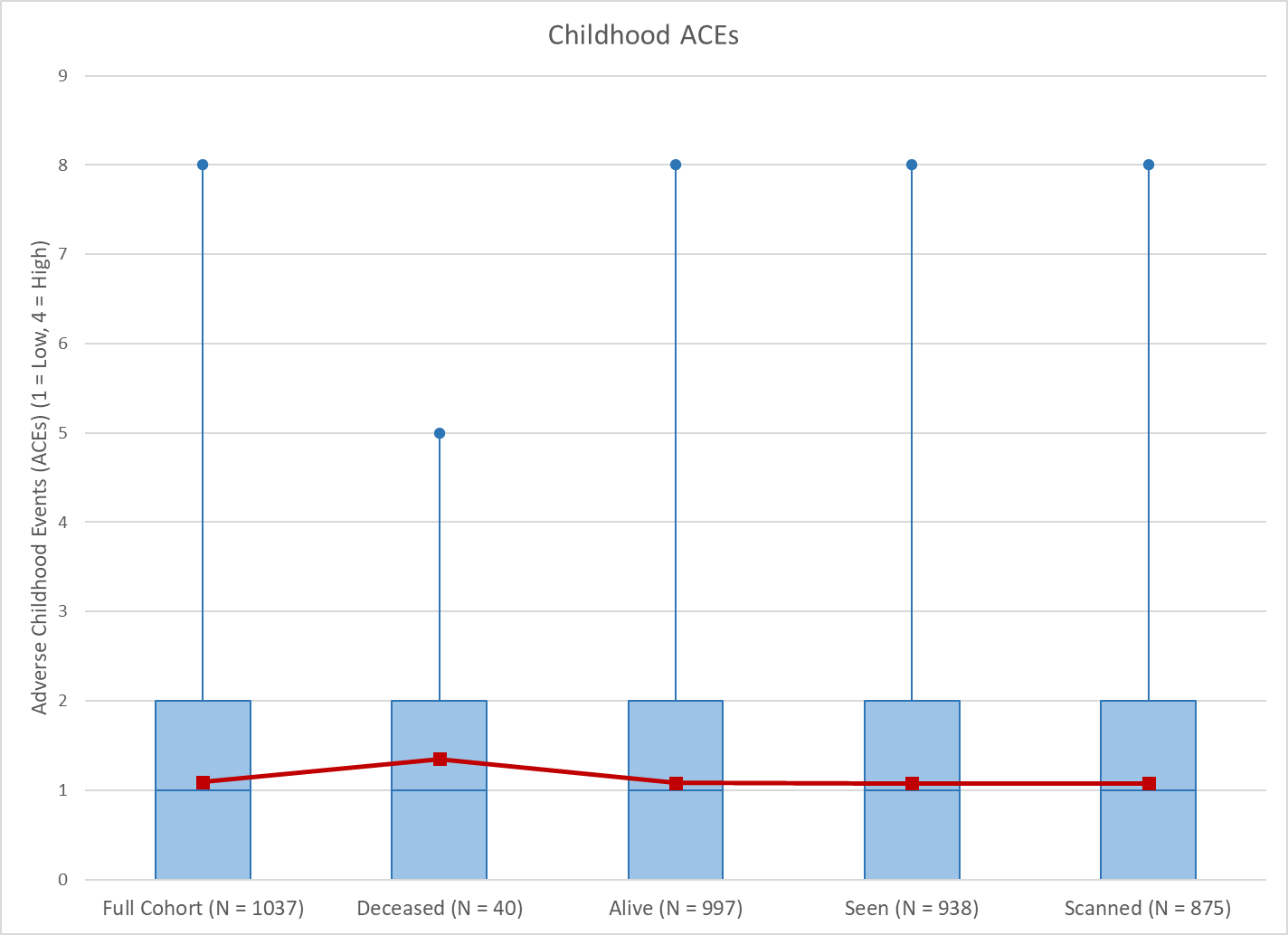

No significant differences were found between the full cohort, those deceased, those alive, those seen at Phase 45 or those scanned at Phase 45 on Childhood Low Self-Control.

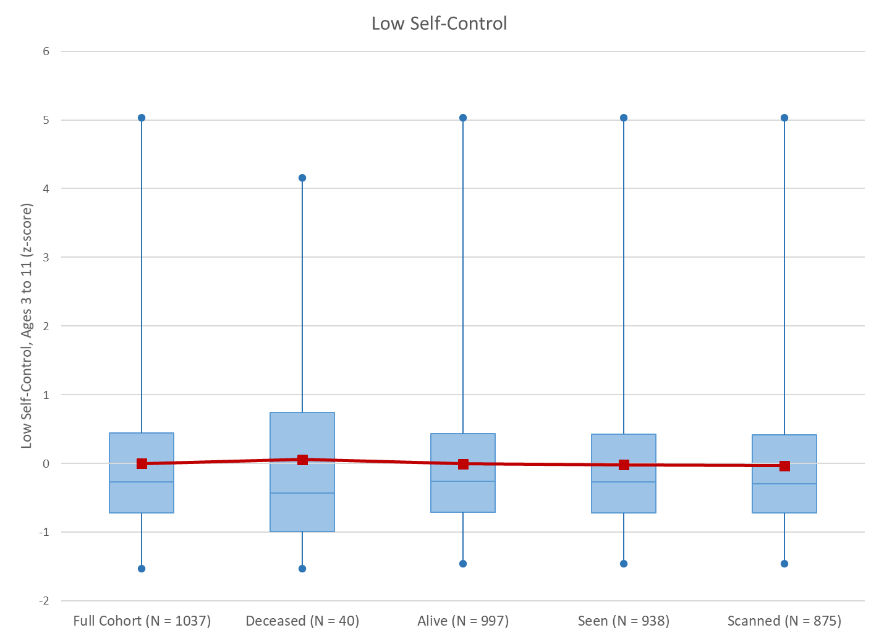

We began collecting DNA from Study members at age 26, in 1998. The DNA biobank does not contain DNA from Study members of Māori descent. No significant differences were found between non-Māori participants with DNA, those who subsequently died, those alive, those seen at Phase 45 or those scanned at Phase 45 on the SSGAC 2021 polygenic score for educational attainment.

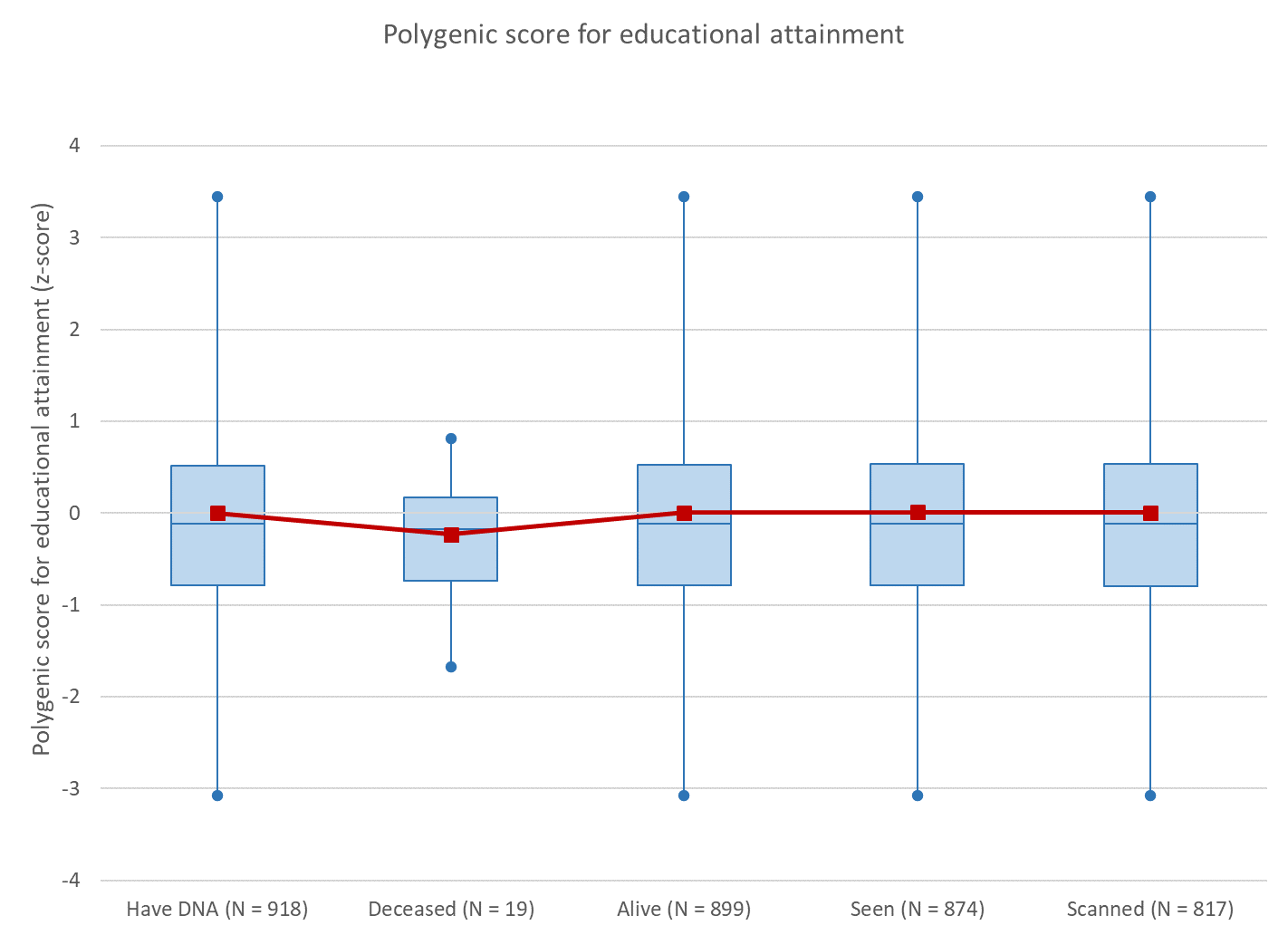

### Magnetic Resonance Imaging

Procedure. Study members were scanned using a MAGNETOM Skyra 3T scanner (Siemens Healthcare, Erlangen, Germany) equipped with a 64-channel head and neck coil (due to head size constraints, seven participants were scanned with a 20-channel head/neck coil) at the Pacific Radiology Group imaging centre in Dunedin, New Zealand, between August 2016 and April 2019. High resolution T1-weighted images, three-dimensional fluid-attenuated inversion recovery (FLAIR) images, and a gradient echo field map were obtained. Structural MRI data were analysed using the Human Connectome Project (HCP) minimal preprocessing pipeline.^1^ Outputs of the preprocessing pipeline were visually checked for accurate surface generation by examining each participant’s myelin map, pial surface, and white matter boundaries. Study personnel who processed the MRI images were masked to participants’ retinal measurements. Of the 875 Study members for whom structural MRI data were available, 4 were excluded due to major incidental findings or previous injuries (e.g., large tumours or extensive damage to the brain/skull), 9 due to missing FLAIR or field map scans, and 1 due to poor surface mapping, yielding 861 datasets for analyses. Additionally, white matter hyperintensities measurements were removed from the dataset for 3 SMs due to multiple sclerosis and 6 Study members due to inaccurate labelling or low-quality data, yielding 852 datasets for WMH analysis.

Image acquisition parameters. High resolution T1-weighted images were obtained using an MP-RAGE sequence with the following parameters: TR=2400 ms; TE=1.98 ms; 208 sagittal slices; flip angle, 9°; FOV, 224mm; matrix=256×256; slice thickness=0.9mm with no gap (voxel size 0.9×0.875×0.875mm); and total scan time=6 min 52 sec. 3D fluid-attenuated inversion recovery (FLAIR) images were obtained with the following parameters: TR=8000ms; TE=399ms; 160 sagittal slices; FOV=240mm; matrix=232×256; slice thickness=1.2mm (voxel size 1.2x0.9×0.9mm); and total scan time=5 min 38 sec. Additionally, a gradient echo field map was acquired with the following parameters: TR=712ms; TE=4.92 and 7.38ms; 72 axial slices; FOV=200mm; matrix=100×100; slice thickness=2.0mm (voxel size 2mm isotropic); and total scan time=2 min 25 sec.

Image processing. Structural MRI data were analyzed using the Human Connectome Project (HCP) minimal preprocessing pipeline as extensively detailed elsewhere.^1^ Briefly, T1-weighted and FLAIR images were processed through the PreFreeSurfer, FreeSurfer, and PostFreeSurfer pipelines. T1-weighted and FLAIR images were corrected for readout distortion using the gradient echo field map, coregistered, brain-extracted, and aligned together in the native T1 space using boundary-based registration.^2^ Images were then processed with a custom FreeSurfer recon-all pipeline that is optimized for structural MRI with higher resolution than 1 mm isotropic. Finally, recon-all output were converted into CIFTI format and registered to common 32k_FS_LR mesh using MSM-sulc.^3^

Brain Age Gap Estimate. The brain Age Gap Estimate (brainAGE; ICC for test-retest reliability=.81) is a score that represents the difference, or gap, between a person’s chronological age and their estimated age based on multiple measures of brain structure including cortical thickness, surface area, and volume of subcortical grey matter, white matter, and cerebrospinal fluid.^4^ The formation of this measure in this cohort has been described previously.^5^

White matter hyperintensities. To identify and extract the total volume of white matter hyperintensities (WMH), T1-weighted and FLAIR images for each participant were processed with UBO Detector, a cluster-based, fully-automated pipeline with high reliability in our data (test-retest ICC=0.87, 95% CI .73-.95) and out of sample performance.^6^ The resulting WMH probability maps were thresholded at 0.7, which is the suggested standard. WMH volume is measured in Montreal Neurological Institute (MNI) space, removing the influence of differences in brain volume and intracranial volume on WMH volume. Because of the potential for bias and false positives due to the thresholds and masks applied in UBO, the resulting WMH maps for each participant were manually checked by two independent raters to ensure that false detections did not substantially contribute to estimates of WMH volume. Visual inspections were done blind to the participants’ cognitive status. Due to the tendency of automated algorithms to mislabel regions surrounding the septum as white matter hyperintensities, these regions were manually masked out, to further ensure the most accurate grading possible.

Subcortical grey matter volume. Grey matter volumes were extracted for 10 subcortical structures using the FreeSurfer aseg parcellation (https://surfer.nmr.mgh.harvard.edu/).

Parcel-wise cortical surface area and cortical thickness. For each subject the mean cortical thickness and surface area were extracted from each of the 360 cortical areas in the HCP-MPP1.0 parcellation.^4^ Subcortical volumes were extracted separately using the automatic segmentation (“aseg”) step of FreeSurfer version 6.0. FreeSurfer version 6.0 was used because the HCP FreeSurfer pipeline was optimized for the cortical surface, resulting in lower-quality segmentation of subcortical volumes in our dataset. Outputs of the minimal preprocessing pipeline were visually checked for accurate surface generation by examining each subject’s myelin map, pial surface, and white matter boundaries. Accuracy of subcortical segmentation was confirmed by visual inspection of the "aseg" labels overlaid on the volumes.

All measures were checked for test-retest reliability (brainAGE ICC=.81; white matter hyperintensities volume ICC=.87; subcortical grey matter volume mean ICC=.956; total cortical surface area ICC=.996; average cortical thickness ICC=.94; parcel-wise cortical thickness mean ICC=.846; parcel-wise surface area mean ICC=.942).

Fractional anisotropy. Diffusion images were processed in FSL (http://fsl.fmrib.ox.ac.uk/fsl). Raw diffusion-weighted images were corrected for susceptibility artifacts, subject movement, and eddy currents using topup and eddy. Images were then skull-stripped and fitted with diffusion tensor models at each voxel using FMRIB's Diffusion Toolbox (FDT; http://fsl.fmrib.ox.ac.uk/fsl/fslwiki/FDT). The resulting FA images from all subjects were non-linearly registered to the FA template developed by the Enhancing Neuro Imaging Genetics Through Meta-Analysis consortium (ENIGMA), a minimal deformation target calculated across a large number of individuals.^7^ The images were then processed using the tract-based spatial statistics (TBSS) analytic method^8^ modified to project individual FA values onto the ENIGMA-DTI skeleton. Following the extraction of the skeletonized white matter and projection of individual FA values, average FA across the full skeleton and average FA within tract-wise regions of interest from the intersection of the skeleton and the 27 regions in the Johns Hopkins University (JHU) white matter parcellation atlas^9^ were calculated. After visual inspection of all diffusion images, 7 study members were removed because data was collected with 20 channel head coils leading to poor diffusion image quality, 3 were removed due to major incidental findings, 5 were removed due to excessive (>3mm) motion detected with the eddy tool, and 6 were removed due to missing diffusion scans. This resulted in 854 study members with quality diffusion images that were included in the analysis. Test-retest reliability, as assessed in 20 Dunedin Study members (mean interval between scans = 79 days), was excellent (ICC=.956 for whole brain average FA, and mean ICC=.879 across 27 tracts).

### Supplementary Tables

### Supplemental table 1. Associations between pTau181 levels at age 45 and cognitive performance.

|  | **Model 1 (pTau181 + sex)** | | | | **Model 2 (pTau181 + sex + BMI)** | | | | **Model 3 (pTau181 + sex + APOEε4)** | | | |
| --- | --- | --- | --- | --- | --- | --- | --- | --- | --- | --- | --- | --- |
|  | **β** | **Lower CI** | **Upper CI** | **p** | **β** | **Lower CI** | **Upper CI** | **p** | **β** | **Lower CI** | **Upper CI** | **p** |
| **Adult IQ** | 0.03 | -0.04 | 0.10 | 0.433 | 0.03 | -0.04 | 0.10 | 0.429 | 0.04 | -0.03 | 0.11 | 0.261 |
| **Child IQ** | 0.01 | -0.05 | 0.08 | 0.733 | 0.01 | -0.05 | 0.08 | 0.726 | 0.03 | -0.03 | 0.10 | 0.336 |
| **Working memory** | 0.04 | -0.03 | 0.10 | 0.299 | 0.03 | -0.03 | 0.10 | 0.314 | 0.04 | -0.03 | 0.11 | 0.242 |
| **Processing speed** | 0.02 | -0.05 | 0.09 | 0.548 | 0.02 | -0.05 | 0.09 | 0.543 | 0.04 | -0.03 | 0.11 | 0.298 |
| **Perceptual reasoning** | 0.01 | -0.06 | 0.08 | 0.797 | 0.01 | -0.06 | 0.08 | 0.800 | 0.01 | -0.06 | 0.09 | 0.694 |
| **Verbal comprehension** | 0.03 | -0.03 | 0.10 | 0.328 | 0.03 | -0.03 | 0.10 | 0.329 | 0.05 | -0.02 | 0.13 | 0.136 |

### Supplemental table 2. pTau181 and global MRI variables (relative values, controlled for total brain volume).

|  | **Model 1 (pTau181 +TBV + sex)** | | | | **Model 2 (pTau181 + TBV + sex + BMI)** | | | | **Model 3 (pTau181 + TBV + sex + APOEε4)** | | | |
| --- | --- | --- | --- | --- | --- | --- | --- | --- | --- | --- | --- | --- |
|  | **β** | **Lower CI** | **Upper CI** | **p** | **β** | **Lower CI** | **Upper CI** | **p** | **β** | **Lower CI** | **Upper CI** | **p** |
| **brainAGE** | -0.04 | -0.10 | 0.03 | 0.290 | -0.04 | -0.10 | 0.03 | 0.285 | -0.06 | -0.13 | 0.01 | 0.108 |
| **Total SA** | -0.01 | -0.03 | 0.02 | 0.674 | -0.01 | -0.03 | 0.02 | 0.691 | -0.01 | -0.04 | 0.02 | 0.664 |
| **Mean CT** | 0.00 | -0.07 | 0.07 | 0.943 | 0.00 | -0.07 | 0.07 | 0.970 | 0.03 | -0.04 | 0.11 | 0.364 |
| **WMH vol (log)** | 0.01 | -0.06 | 0.08 | 0.847 | 0.01 | -0.06 | 0.08 | 0.843 | 0.01 | -0.06 | 0.09 | 0.699 |
| **Mean FA** | -0.00 | -0.07 | 0.06 | 0.911 | -0.00 | -0.07 | 0.06 | 0.933 | 0.03 | -0.04 | 0.09 | 0.433 |

*Note.* TBV: total brain volume; BMI: body mass index; SA: surface area; CT: cortical thickness, WMH vol: white matter hyperintensity volume (log transformed); FA: fractional anisotropy.

### Supplemental table 3. pTau181 and grey matter volumes of subcortical structures (absolute values).

|  | **Model 1 (pTau181 + sex)** | | | | **Model 2 (pTau181 + sex + BMI)** | | | | **Model 3 (pTau181 + sex + APOEε4)** | | | |
| --- | --- | --- | --- | --- | --- | --- | --- | --- | --- | --- | --- | --- |
|  | **β** | **Lower CI** | **Upper CI** | **p** | **β** | **Lower CI** | **Upper CI** | **p** | **β** | **Lower CI** | **Upper CI** | **p** |
| **Accumbens** | 0.00 | -0.06 | 0.07 | 0.893 | 0.01 | -0.06 | 0.07 | 0.877 | 0.02 | -0.05 | 0.09 | 0.498 |
| **Amygdala** | 0.04 | -0.02 | 0.10 | 0.229 | 0.04 | -0.02 | 0.10 | 0.224 | 0.05 | -0.01 | 0.12 | 0.087 |
| **Caudate** | -0.02 | -0.09 | 0.04 | 0.512 | -0.02 | -0.09 | 0.04 | 0.517 | -0.00 | -0.07 | 0.07 | 0.936 |
| **Cerebellum** | 0.03 | -0.03 | 0.10 | 0.300 | 0.03 | -0.03 | 0.09 | 0.315 | 0.05 | -0.01 | 0.12 | 0.115 |
| **Hippocampus** | -0.02 | -0.09 | 0.04 | 0.461 | -0.02 | -0.09 | 0.04 | 0.472 | -0.00 | -0.07 | 0.06 | 0.984 |
| **Pallidum** | 0.03 | -0.03 | 0.09 | 0.354 | 0.03 | -0.03 | 0.09 | 0.363 | 0.04 | -0.03 | 0.10 | 0.242 |
| **Putamen** | 0.03 | -0.03 | 0.09 | 0.317 | 0.03 | -0.03 | 0.09 | 0.330 | 0.04 | -0.02 | 0.11 | 0.177 |
| **Thalamus** | -0.01 | -0.07 | 0.05 | 0.766 | -0.01 | -0.07 | 0.05 | 0.770 | 0.01 | -0.05 | 0.08 | 0.655 |
| **Ventral diencephalon** | 0.00 | -0.06 | 0.06 | 0.886 | 0.00 | -0.06 | 0.06 | 0.905 | 0.03 | -0.03 | 0.09 | 0.383 |
| **Brain stem** | -0.01 | -0.07 | 0.06 | 0.843 | -0.01 | -0.07 | 0.05 | 0.818 | 0.01 | -0.06 | 0.07 | 0.790 |

### Supplemental table 4. pTau181 and grey matter volumes of subcortical structures (relative values, controlled for total brain volume)

|  | **Model 1 (pTau181 +TBV + sex)** | | | | **Model 2 (pTau181 + TBV + sex + BMI)** | | | | **Model 3 (pTau181 + TBV + sex + APOEε4)** | | | |
| --- | --- | --- | --- | --- | --- | --- | --- | --- | --- | --- | --- | --- |
|  | **β** | **Lower CI** | **Upper CI** | **p** | **β** | **Lower CI** | **Upper CI** | **p** | **β** | **Lower CI** | **Upper CI** | **p** |
| **Accumbens** | -0.01 | -0.06 | 0.05 | 0.823 | -0.01 | -0.06 | 0.05 | 0.848 | -0.00 | -0.06 | 0.06 | 0.929 |
| **Amygdala** | 0.03 | -0.02 | 0.07 | 0.307 | 0.03 | -0.02 | 0.08 | 0.295 | 0.03 | -0.02 | 0.08 | 0.296 |
| **Caudate** | -0.03 | -0.09 | 0.02 | 0.249 | -0.03 | -0.09 | 0.02 | 0.255 | -0.03 | -0.09 | 0.03 | 0.336 |
| **Cerebellum** | 0.02 | -0.03 | 0.08 | 0.407 | 0.02 | -0.03 | 0.07 | 0.425 | 0.03 | -0.03 | 0.09 | 0.330 |
| **Hippocampus** | -0.03 | -0.09 | 0.02 | 0.182 | -0.03 | -0.09 | 0.02 | 0.192 | -0.03 | -0.08 | 0.03 | 0.301 |
| **Pallidum** | 0.02 | -0.03 | 0.07 | 0.498 | 0.02 | -0.03 | 0.07 | 0.504 | 0.01 | -0.04 | 0.06 | 0.748 |
| **Putamen** | 0.02 | -0.03 | 0.07 | 0.436 | 0.02 | -0.03 | 0.07 | 0.449 | 0.02 | -0.04 | 0.07 | 0.520 |
| **Thalamus** | -0.02 | -0.06 | 0.02 | 0.227 | -0.02 | -0.06 | 0.02 | 0.233 | -0.02 | -0.06 | 0.02 | 0.294 |
| **Ventral diencephalon** | -0.01 | -0.05 | 0.03 | 0.650 | -0.01 | -0.05 | 0.03 | 0.635 | -0.01 | -0.05 | 0.04 | 0.794 |
| **Brain stem** | -0.02 | -0.07 | 0.03 | 0.386 | -0.02 | -0.07 | 0.02 | 0.369 | -0.02 | -0.07 | 0.02 | 0.320 |

*Note.* TBV: total brain volume; BMI: body mass index.
